# Early mineralocorticoid receptor antagonism in diabetic nephropathy limits albuminuria by preserving the glomerular endothelial glycocalyx

**DOI:** 10.1101/2021.05.13.21252519

**Authors:** Michael Crompton, Joanne K. Ferguson, Raina D. Ramnath, Karen L. Onions, Anna S. Ogier, Colin J. Down, Laura Skinner, Lauren K. Dixon, Judit Sutak, Steven J. Harper, Paola Pontrelli, Loreto Gesualdo, Gavin I. Welsh, Rebecca R. Foster, Simon C. Satchell, Matthew J. Butler

## Abstract

The glomerular endothelial glycocalyx (GEnGlx) forms the first part of the glomerular filtration barrier (GFB). We have previously shown that excess mineralocorticoid receptor (MR) activation causes GEnGlx damage and albuminuria. Damage to the GEnGlx occurs early in the pathogenesis of diabetic nephropathy (DN). Here we sought to determine whether MR antagonism with spironolactone could prevent the development of albuminuria in diabetes, by preserving the GEnGlx to maintain the GFB. Streptozotocin-induced diabetic Wistar rats developed increased glomerular albumin permeability (P*s’*_*alb*_) and albuminuria, with associated GEnGlx loss and increases in plasma and urine active matrix metalloproteinase 2 (MMP2). MR antagonism reduced urine active MMP2, preserved the GEnGlx, restored P*s’*_*alb*_ to control values and prevented diabetes-induced albuminuria progression. Enzymatic degradation of the GEnGlx, with hyaluronidase, reversed the effect of MR antagonism in diabetic rats, confirming the importance of GEnGlx preservation in this model. Using this model we validated a novel fluorescent profile peak-to-peak confocal imaging technique and applied it to assess GEnGlx damage on renal biopsies from patients with DN and compared them to healthy controls. We confirmed that GEnGlx loss occurs in human DN and may contribute to the disease phenotype. Taken together our work suggests GEnGlx preservation as an important novel mechanism for reno-protection by MR inhibitors in diabetes.

**Translational Statement:** Mineralocorticoid receptor (MR) antagonists reduce albuminuria in diabetic nephropathy (DN), but side effects limit their clinical utility, and the mechanism is unknown. This paper demonstrates that MR antagonism prevented the development of glomerular endothelial glycocalyx (GEnGlx) dysfunction and albuminuria. Highly specific, enzyme mediated, removal of the GEnGlx abolished this effect, confirming the importance of EnGlx preservation in this model. Our data also confirm that GEnGlx damage occurs in human DN and may contribute to the clinical phenotype. This work suggests directly targeting glycocalyx preservation in DN could reproduce the protective effects seen with MR inhibition, whilst avoiding side effects.

## Introduction

Glomerular diseases, including diabetic nephropathy (DN), are the commonest cause of end-stage renal failure.^1^ DN is a serious complication of diabetes, with approximately 1 in 5 people with diabetes needing treatment for DN during their lifetime.^2^ In the UK, DN alone accounts for ∼11,000 people requiring renal replacement therapy (RRT) and 18% of all people requiring RRT do so because of diabetes.^1^

Renin-Angiotensin-Aldosterone System (RAAS) blockade with both angiotensin-converting enzyme inhibitors (ACEi) or angiotensin-receptor blockers (ARB) reduce albuminuria and the risk of end-stage kidney disease.^3^ However, because of aldosterone escape, blockade of RAAS with ACEi or ARB may not be effective at suppressing aldosterone-induced mineralocorticoid receptor (MR) stimulation.^4,5^ In DN patients taking ACEi or ARB, the addition of MR antagonists further reduces albuminuria, indicating that MR activation contributes to albuminuria.^6–11^ Most recently, the large Phase III FIDELIO-DKD trial suggests that in patients with chronic kidney disease (CKD) and type 2 diabetes, MR inhibition reduced the risk of CKD progression and cardiovascular events.^12^ However, side effects, such as hyperkalaemia, limit the clinical use of MR antagonists.^9,11,13–15^ Thus, better definition of mechanisms of glomerular damage are needed to identify novel tissue-specific therapeutic targets.

Changes in the glomerular endothelium are increasingly recognised in DN and other glomerular diseases.^16^ The glomerulus is the filtering unit of the kidney and its function is dependent on the multi-layer structure of the glomerular filtration barrier (GFB). This consists of glomerular endothelial cells (GEnC), the glomerular basement membrane (GBM) and podocytes.^17^ The glomerular endothelial glycocalyx (GEnGlx) covers the luminal surface of the GEnC, filling the fenestrations, contributing to glomerular barrier function.^18,19^ Our group and others have shown that GEnGlx specifically limits albumin permeability *in vitro*^20–22^ and *in vivo*^23–26^. The EnGlx is a hydrated poly-anionic gel composed principally of proteoglycan core proteins, glycosaminoglycan chains and sialoglycoproteins.^27^ In healthy vascular physiology the EnGlx has multiple roles, including regulating vascular permeability,^28,29^ mediating shear stress mechanotransduction,^30,31^ and attenuating immune cell–endothelium interactions,^32,33^ with further roles under investigation.^27,34,35^ Disruption of the EnGlx has been linked to the development of various clinical conditions including; diabetes, sepsis, preeclampsia, and atherosclerosis.^18,27,34,36^

In humans, DN is characterised by albuminuria.^18,37^ In early DN no macroscopic GBM or podocyte changes are detectable, but systemic endothelial and glycocalyx dysfunction in both type 1^38^ and type 2 diabetes^39^ has been shown to occur. Others have previously shown glycosaminoglycans are lost from the GFB in diabetes,^40,41^ and we have confirmed GEnGlx loss in diabetic mice^42,43^ and rats.^26^ Together these findings strongly implicate GEnGlx damage as a key initiator of albuminuria in DN.^18,19,37^

EnGlx components are cleaved from the cell surface by “sheddases”, including matrix metalloproteinases (MMPs).^44,45^ We have identified MMP2 and MMP9 as key glycocalyx sheddases.^43,45,46^ We have recently defined a pathway whereby excess MR activation results in increased MMP2 and MMP9 activity and consequent GEnGlx dysfunction and albuminuria.^46^ Here we seek to determine whether this pathological pathway is responsible for GEnGlx damage in diabetes.

We hypothesize that MR antagonism reduces MMP activity in diabetes, preserving the GEnGlx and limiting the development of albuminuria.

## Methods

### Animals

All animal protocols were approved by the UK Government Home Office and conformed to the National Institutes of Health Guide for the Care and Use of Laboratory Animals. Male Wistar rats (150–200g) were purchased from Charles River UK Limited (Kent, UK) and maintained by the Animal Services Unit, University of Bristol. Animals were housed in a conventional facility with a controlled environment (21–24 °C and 12:12 hour light–dark cycle). A period of 1 to 2 weeks was allowed for animals to acclimatize before any experiments. All rats had free access to food and water throughout the study. Timelines of the experimental protocols used are included (Figure 1A and 4A).

**Figure 1.**
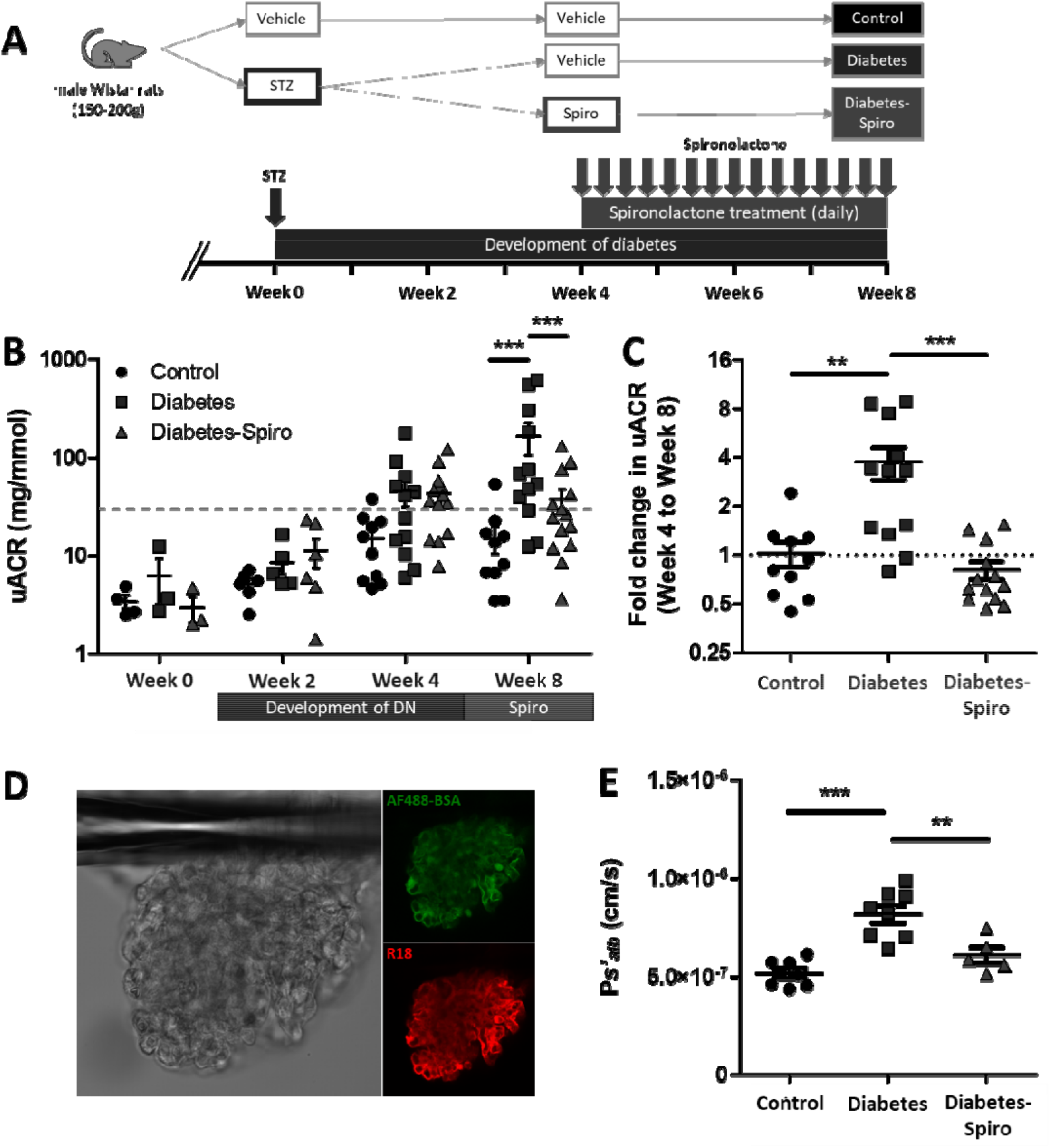
Development of albuminuria and increased glomerular permeability in early diabetic nephropathy is ameliorated by MR antagonism with spironolactone. (A) Schematic overview of streptozotocin (STZ)-induced diabetic model and spironolactone (spiro) treatment protocol for male Wistar rats. An injection of STZ was given at week 0. Four weeks post-STZ injection, spiro (an MR inhibitor) was given for 28 days and rats were culled at week 8 post-STZ injection. Rats were randomised to receive STZ and spiro. (B) Albuminuria levels stabilise in diabetic rats treated with spiro but progressively worsen in rats given vehicle. Urinary albumin:creatinine ratio (uACR) was determined at week 0 (control, *n*=4; diabetes, *n*=3; diabetes-spiro, *n*=3), week 2 (control, *n*=6; diabetes, *n*=5; diabetes-spiro, *n*=6), week 4 (control, *n*=10; diabetes, *n*=12; diabetes-spiro, *n*=13), and week 8 (control, *n*=10; diabetes, *n*=12; diabetes-spiro, *n*=14). A base-10 log scale is used for the Y axis. (C) Treatment with spiro for 28 days reduced the fold change in uACR from initiation of treatment, week 4 to week 8 (control, *n*=10; diabetes, *n*=12; diabetes-spiro, *n*=13). A base-2 log scale is used for the Y axis. (D) Representative images of an isolated glomerulus stained with R18 and Alexa Fluor 488–bovine serum albumin (AF488-BSA). (E) Glomerular albumin permeability (P*s’*_*alb*_) was measured at week 8 (control, *n*=7 rats (32 glomeruli); diabetes, *n*=8(36); diabetes-spiro, *n*=5(21)). Each dot, triangle, and square on the graph represents a rat. Data are expressed as mean ± SEM. ** *P*<0.01; *** *P*<0.001.

### Human Renal Samples

All studies on human kidney tissue were approved by national and local research ethics committees (REC) and conducted in accordance with the tenets of the Declaration of Helsinki. Renal biopsy samples from 34 patients were obtained from Bristol, UK (Histopathology Department, Southmead Hospital) and Bari, Italy (Division of Nephrology, Dialysis and Transplantation, Department of Emergency and Organ Transplantation, Aldo Moro University of Bari). Samples provided in Bristol were archived anonymised samples (REC H0102/45). For Bari samples, patients gave written informed consent for the use of this material for research (Study no. 5156/2017). Tissue was immersion fixed in 10% neutral buffered formalin and embedded in paraffin. Samples had been clinicopathologically diagnosed, including 19 patients with diabetic nephropathy (DN), 8 patients with thin basement membrane nephropathy (TBMN) and 7 histologically normal control patients (Table 1).

**Table 1.** Clinical and analytical data at the time of renal biopsy.

|  | **TBMN -**<br>**Bristol**<br>**(n=8)** | **Control -**<br>**Bari**<br>**(n=7)** | **Diabetes -**<br>**Bristol**<br>**(n=12)** | **Diabetes -**<br>**Bari**<br>**(n=7)** |
| --- | --- | --- | --- | --- |
| Median age<br>(range) | 52<br>(31-73) | 37<br>(8-57) | 51<br>(25 to 78) | 62<br>(43 to 78) |
| Male | 2 | 5 | 6 | 5 |
| Female | 6 | 2 | 6 | 2 |
| DM Type 2 / Type 1 | - | - | 9/3 | 7/0 |
| **Medication** |  |  |  |  |
| Taking ACEi/ARB | 1/8 | 1/7 | 7/12 | 5/7 |
| Taking MR inhibitors | 0/8 | 0/7 | 0/12 | 0/7 |
| Median number of diabetic<br>medications (range) | - | - | 2<br>(1-3) | 2<br>(1-3) |
| **Clinical Results** |  |  |  |  |
| eGFR Median<br>(range) | 70<br>(26-113) | 105<br>(82-128) | 32<br>(18-64) | 22<br>8-82 |
| Median serum albumin, g/L<br>(range) | 36<br>(33-47) | - | 24<br>(14-37) | 30<br>(26-43) |
| Median uPCR, mg/mmol<br>(range) | 21<br>(0-54) | - | 624<br>(12-1416) | 381<br>(16-681) |
| Median uACR, mg/mmol<br>(range) | 0<br>(0-49) | - | 460<br>(0-1279) | 360<br>(68-681) |

TBMN, thin basement membrane nephropathy; DM, diabetes mellitus; ACEi, angiotensin-converting enzyme inhibitors; ARB, angiotensin receptor blocker; MR, mineralocorticoid receptor; eGFR, estimated glomerular filtration rate; uPCR, urinary protein:creatinine ratio; uACR, urinary albumin:creatinine ratio.

TBMN is the most common cause of persistent hematuria and patients usually display minimal or no proteinuria, normal renal function, and a uniformly thinned GBM.^47^ TBMN occurs in more than 1% of the population and is a lifelong non-progressive disorder.^48^ With limited numbers of histologically normal renal biopsy samples, collected in a comparable way to disease samples, renal biopsy samples from TBMN patients represent a good alternative for comparison.

### Fluorescent profile confocal peak-to-peak analysis

Initially, the fluorescent profile peak-to-peak assessment was carried out, blinded with our manual methodology, as previously described.^46,49^ A perpendicular profile line (ROI in Figure 2Ai) was drawn from the inside to the outside of the capillary loop crossing the lectin-labelled glycocalyx first followed by the R18 labelled endothelial membrane. Fluorescence intensity profiles were then generated for the lectin-labelled components of the endothelial glycocalyx and endothelial cell label. The distance between the peak signals from the lectin-488 and the R18 labels (peak-to-peak) is an index of glycocalyx thickness (Figure 2B). Mean data was determined from an average of 3 lines per capillary loop, 3 loops per glomerulus, 3 glomeruli per rat.

**Figure 2.**
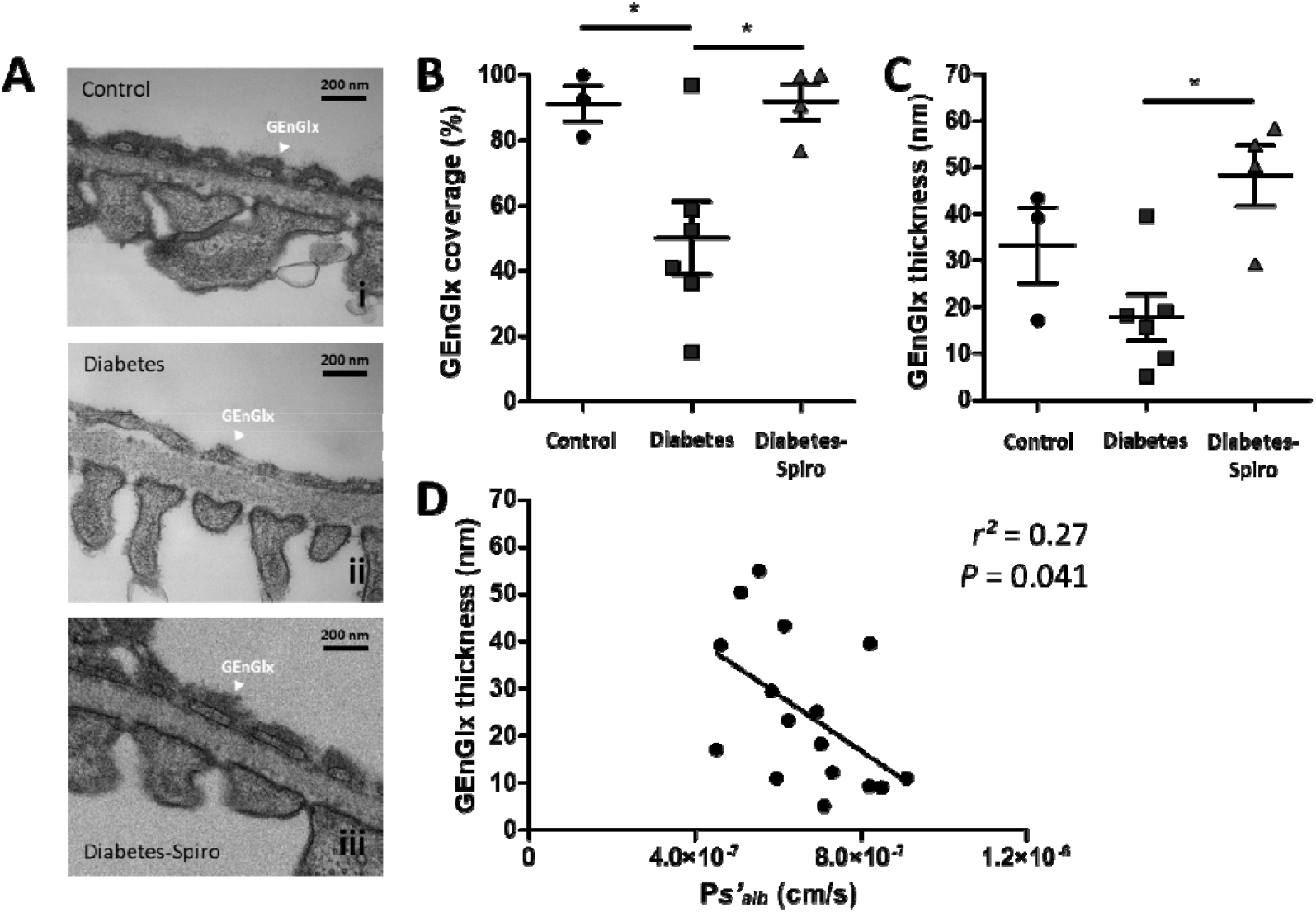
Transmission electron microscopy demonstrated that MR antagonism prevented diabetes-induced glomerular endothelial glycocalyx damage. Rats were perfusion-fixed for transmission electron microscopy (TEM) with cacodylate buffer containing glutaraldehyde and Alcian blue. (A) Representative electron micrographs of the glomerular capillary wall are shown for (i) control, (ii) diabetes, and (iii) diabetes-spironolactone (spiro) samples. Labels indicate glomerular endothelial glycocalyx (GEnGlx). Bars = 200 nm. Quantification at week 8 post-STZ of GEnGlx coverage, and (C) GEnGlx thickness (control, *n*=3 rats (9 glomeruli); diabetes, *n*=6(13); diabetes-spiro, *n*=4(8)). (F) The rate of glomerular albumin leakage (P*s’*_*alb*_) is weakly associated with GEnGlx thickness measured by TEM (*n*=16). Each dot, triangle, and square on the graph represents a rat. Data are expressed as mean ± SEM. * *P*<0.05.

Our peak-to-peak measurement technique was subsequently updated to use a blinded automated methodology. This increased the number of measurements taken from each capillary loop and reduced the time taken to analyse each glomerulus. Briefly, we developed an ImageJ macro to take multiple measurements in a pre-selected capillary loop and generate fluorescence intensity profiles for the lectin components of the GEnGlx and endothelial cell label. Gaussian curves were applied to the raw intensity data of each plot for peak-to-peak measurements (dashed lines in Figure 2B). Mean data was determined from 200 lines per capillary loop, 3 loops per glomerulus, 4-6 glomeruli per rat. Data was excluded with a standard deviation >7.5 and/or a signal-to-noise ratio <15

### Statistical analysis

All statistics were calculated using Prism 8 (GraphPad, CA, USA). Normality was assessed visually and using the Shapiro-Wilk test. Where data was normally distributed an ANOVA or Student’s t-test were conducted. If data was not normally distributed the non-parametric equivalents of these tests were used (Kruskal-Wallis One Way Analysis of Variance on Ranks or Mann-Whitney Rank Sum Test) to establish if data were significant. The Tukey method (ANOVA) or Dunn’s method (Ranks) was used for multiple comparisons between groups. All data are expressed as mean +/- SEM (standard error of the mean) unless stated otherwise. Results with values of *p* < 0.05 were considered statistically significant. Individual rats were considered experimental units within this study.

Please see the Supplementary Methods for details on the type 1 diabetic model and dosing regimen, tissue collection, glomerular albumin permeability (Ps’alb) assay, transmission electron microscopy, lectin staining and MMP activity.

## Results

### Development of albuminuria and increased glomerular permeability in early DN is ameliorated by MR antagonism

Male Wistar rats were hyperglycaemic 3 weeks after receiving STZ (Figure S1A). There was no significant change in body weight in the STZ-treated rats when compared at week 0, prior to STZ injection (Figure S1B). However, from week 4 the diabetic rats gained significantly less weight than control rats, resulting in a 1.3-fold lower body weight after week 8 post-STZ (Figure S1B). Treatment with spironolactone had no significant impact on hyperglycaemia, or on body weight (Figure S1B).

STZ-induced diabetic rats progressively developed albuminuria (Figure 1B) and this became significant with a 10.9-fold increase in albuminuria at week 8 post-STZ (Figure 1B). Spironolactone limited progression of albuminuria in diabetic rats. At week 8 post-STZ diabetic rats treated with spironolactone had a significantly reduced uACR compared to vehicle-treated diabetic rats, with no significant difference when compared to controls (Figure 1B). The fold change in uACR from initiation of the treatment phase (week 4), with vehicle or spironolactone, to when the rats were culled (week 8) was determined (Figure 1C). Diabetic rats had a significant increase in fold change of uACR, compared to both controls and spironolactone-treated diabetic rats. There was no significant difference when comparing spironolactone-treated diabetic rats and controls. The fold change in uPCR was also determined to investigate whether changes in albumin excretion reflected overall protein excretion (Figure S1C). Similarly, diabetic rats had a significant increase in fold change of uPCR, compared to both controls and spironolactone-treated diabetic rats.

Our glomerular permeability assay was used to directly measure the albumin permeability (P*s’*_*alb*_) of individually trapped glomeruli.^26^ In contrast to urine-based measurements this assay directly measures the GFB permeability to albumin and is independent of haemodynamic factor and tubular albumin handling (Figure 1D). Compromised glomerular capillary wall integrity was confirmed by a 1.6-fold increase in P*s’*_*alb*_ (Figure 1E). Spironolactone restored P*s’*_*alb*_ to control values with no significant difference when compared to controls, and permeability was significantly reduced compared to diabetic rats (Figure 1E).

### Diabetes-induced GEnGlx damage in early DN is prevented by MR antagonism

Following removal of the left kidney rats were perfusion-fixed with glutaraldehyde and Alcian blue for TEM,^26,42^ and electron micrographs of the glomerular capillary wall were used to measure a range of glomerular parameters (Figure 2A and S2A). STZ-induced diabetic rats had a 1.8-fold decrease in GEnGlx coverage (Figure 2B). Similarly, there was a 1.9-fold decrease in GEnGlx thickness in diabetic rats, however this was not significant (Figure 2C). Spironolactone treatment in diabetic rats restored both the GEnGlx coverage and thickness (Figure 2B and 2C). Diabetic rats treated with spironolactone had significantly increased GEnGlx coverage and GEnGlx thickness compared to vehicle-treated diabetic rats, with no significant differences when compared to controls (Figure 2B and 2C). Glomerular permeability changes were weakly associated with GEnGlx thickness measured by EM (Figure 2D), suggesting EM based measurements provide a poor measure of glycocalyx integrity. Finally, there were no significant changes in other ultrastructural features, including podocyte glycocalyx (pGlx) thickness, pGlx coverage, GBM width, fenestration density, podocyte foot process density, podocyte foot process width, and podocyte slit diaphragm width (Figure S2B-H).

*Marasmium oreades* agglutinin (MOA) and wheat germ agglutinin (WGA) lectin both bound to the EnGlx on the luminal surface of the labelled GEnC membrane (Figure 3A). We analysed the fluorescent images generated using a fluorescent profile peak-to-peak measurement technique^43,46,49^ to provide an index of GEnGlx thickness (Figure 3B). Manual peak-to-peak measurement of MOA labelling demonstrated a 1.5-fold reduction in GEnGlx thickness in diabetic rats (Figure 3C). Spironolactone treatment in diabetic rats restored the GEnGlx thickness, with no significant differences compared to controls (Figure 3C). Glomerular albumin permeability corelated inversely with the GEnGlx thickness measured by peak-to-peak analysis of MOA/R18 labelling (Figure 3D).

**Figure 3.**
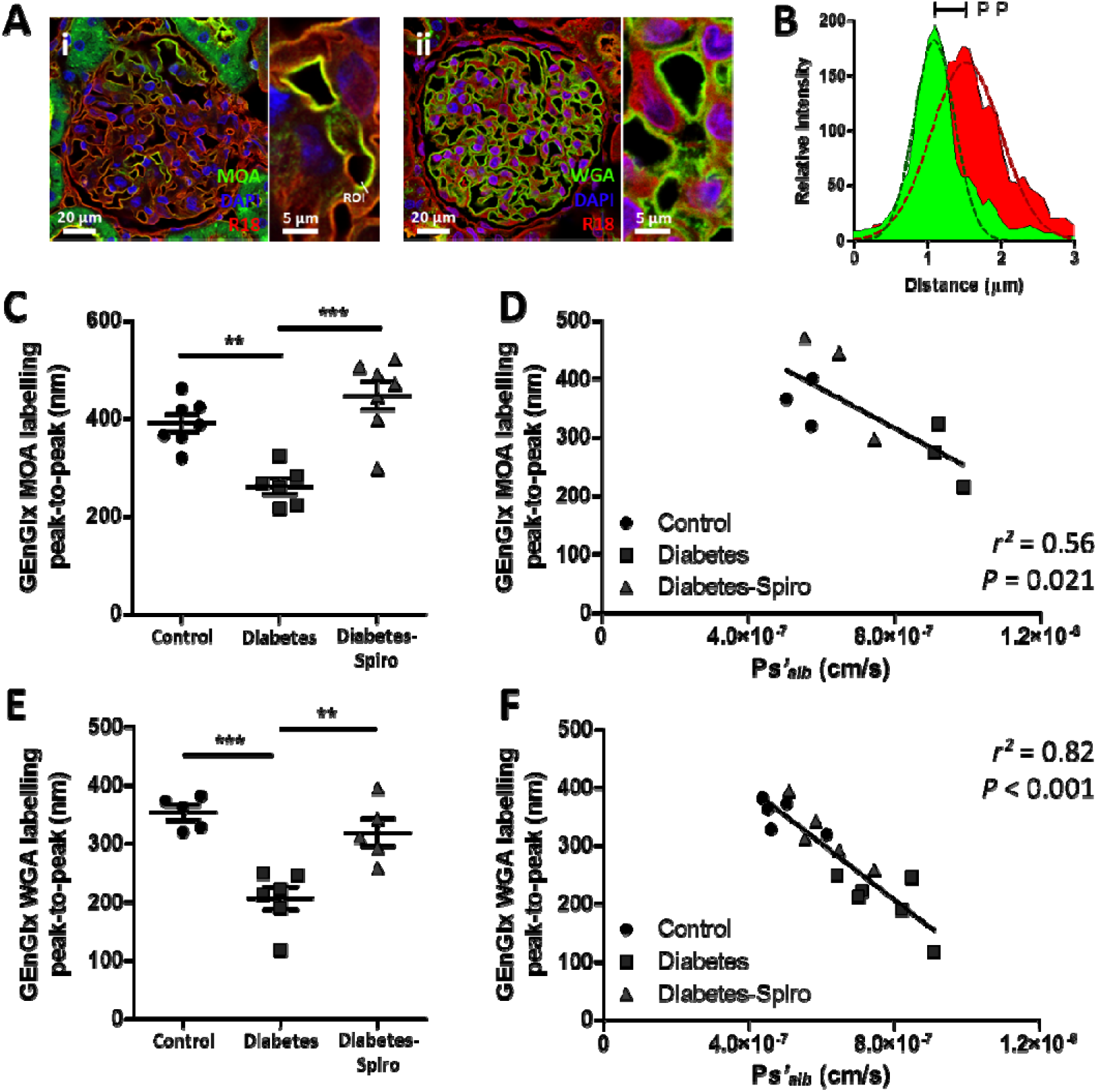
Fluorescent profile peak-to-peak measurements confirm that glomerular endothelial glycocalyx damage is prevented by MR antagonism and correlate strongly with glomerular albumin permeability. (A) Representative images show glomerular capillaries labelled red (R18) and the luminal glomerular endothelial glycocalyx (GEnGlx) labelled green with (i) *Marasmium oreades* agglutinin (MOA) or (ii) wheat germ agglutinin (WGA). Bars = 20 _μ_m and 5 _μ_m. ROI, region of interest for fluorescent profile peak-to-peak measurement. (B) Representative relative intensity peaks of R18 (red) and MOA (green) profiles showing peak-to-peak (P-P) assessment of the GEnGlx – Gaussian curves (dashed lines) were fit to the raw intensity data of each plot for peak-to-peak measurements. Quantification at week 8 post-STZ of GEnGlx MOA labelling peak-to-peak (control, *n*=7; diabetes, *n*=6; diabetes-spironolactone (spiro), *n*=7) and (D) functional association with the rate of glomerular albumin leakage (P*s’*_*alb*_) (*n*=9). Quantification at week 8 post-STZ of (E) GEnGlx WGA labelling peak-to-peak (control, *n*=5; diabetes, *n*=6; diabetes-spiro, *n*=5) and (F) functional association with the rate of glomerular P*s’*_*alb*_ (*n*=16). Each dot, triangle, and square on the graph represents a rat. Data are expressed as mean ± SEM. ** *P*<0.01; *** *P*<0.001.

To confirm the validity of our findings with MOA labelling we applied a second lectin, WGA, for peak-to-peak measurements, and developed an automated methodology. As with MOA, we found significant glycocalyx damage in diabetic rats, with a 1.7-fold decrease in GEnGlx thickness (Figure 3E). Spironolactone in diabetic rats significantly restored the GEnGlx thickness, with no significant differences compared to controls (Figure 3E). Again, glomerular permeability changes corelated inversely, and strongly, with GEnGlx thickness measured by peak-to-peak of WGA/R18 labelling (Figure 3F), suggesting peak-to-peak provides a superior measure of glycocalyx integrity compared to EM assessment.

### The effect of MR antagonism in preventing the diabetes-induced increase in glomerular permeability is dependent on the GEnGlx

Hyaluronidase, a glycocalyx-degrading enzyme, was infused in a sub-group of spironolactone-treated diabetic rats to remove the EnGlx to confirm the importance of GEnGlx preservation in this model (Figure 4A). Automated peak-to-peak measurement of MOA labelling demonstrated a significant reduction in GEnGlx thickness with enzyme treatment (Figure 4B). Similarly, WGA labelling and analysis showed a significant reduction in thickness with enzyme treatment (Figure 4C). Enzymatic degradation of the GEnGlx significantly increased the uACR by 3.6-fold and returned albuminuria to diabetes-induced levels (Figure 4D). Electron micrographs of perfusion fixed glomeruli confirmed that enzyme treatment resulted in significant GEnGlx loss, evidenced by a 1.5-fold decrease in GEnGlx coverage (Figure 4E and 4F) and a 2.9-fold decrease in GEnGlx thickness (Figure 4G). Enzyme treatment had no significant effect on either pGlx coverage or thickness or GBM width, suggesting that hyaluronidase activity remained focused on the luminal EnGlx (Figure S3).

**Figure 4.**
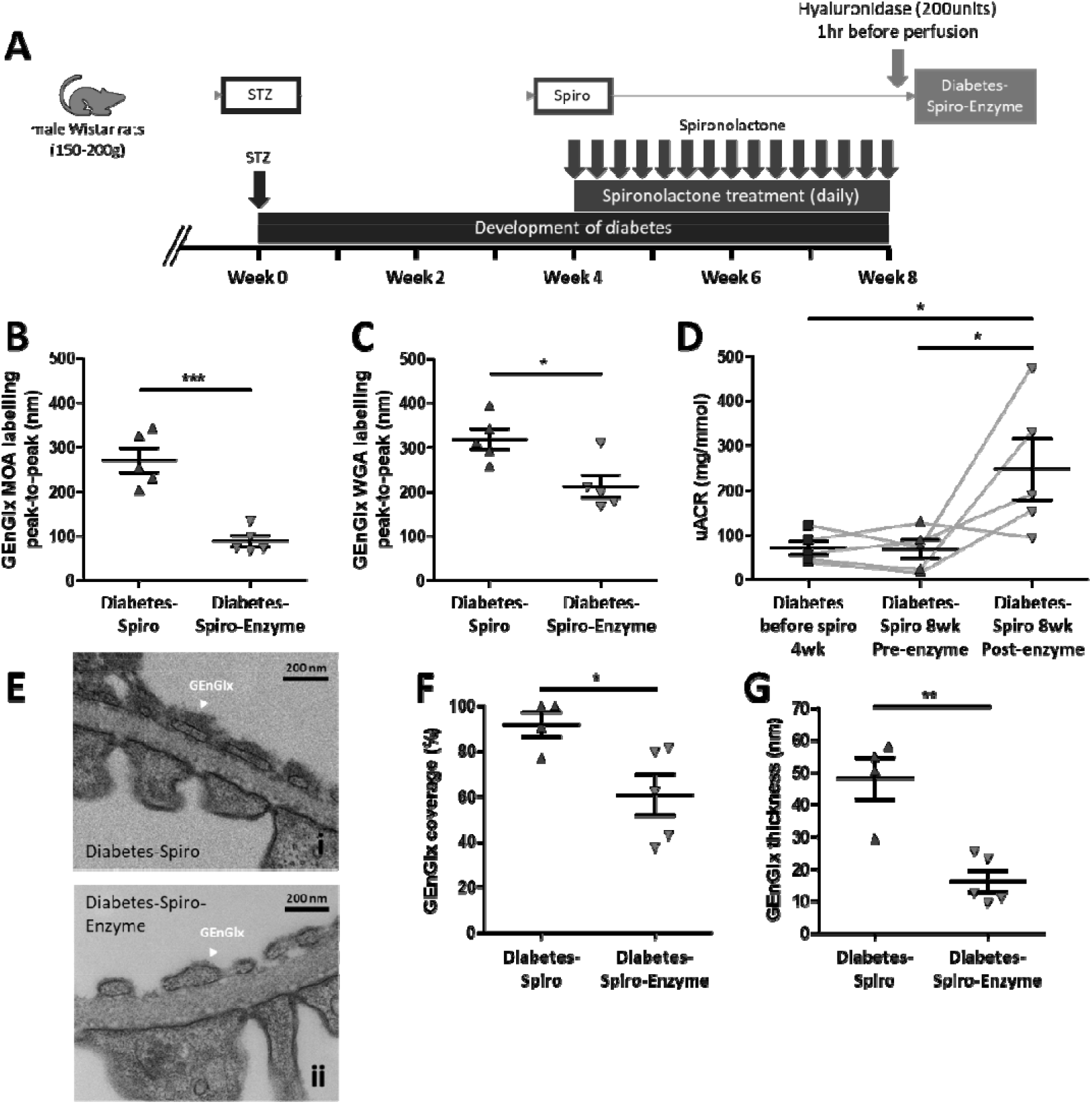
The effect of MR antagonism in preventing the diabetes-induced increase in glomerular permeability is dependent on the GEnGlx. (A) Schematic overview of enzymatic degradation of the glomerular endothelial glycocalyx (GEnGlx) with hyaluronidase on spironolactone (spiro) treated male Wistar rats. An injection of streptozotocin (STZ) was given at week 0. Four weeks post-STZ injection, spiro (an MR inhibitor) was given for 28 days and rats were given hyaluronidase at week 8 post-STZ via tail vein injection 1 hour before being culled for tissue collection. Rats were randomised to receive hyaluronidase. Quantification at week 8 post-STZ of (B) GEnGlx WGA labelling peak-to-peak (diabetes-spiro, *n*=5; diabetes-spiro-enzyme, *n*=5) and (C) GEnGlx MOA labelling peak-to-peak (diabetes-spiro, *n*=5; diabetes-spiro-enzyme, *n*=5) confirmed enzyme degradation of GEnGlx. (D) Albuminuria levels returned to those expected in vehicle treated diabetic rats. Urinary albumin:creatinine ratio (uACR) was determined from the same rats (*n*=5) at week 4 post-STZ (diabetes before spiro 4wk), week 8 post-treatment with spiro (diabetes-spiro 8wk pre-enzyme), and week 8 post-hyaluronidase (diabetes-spiro 8wk post-enzyme). The connecting line (grey) represents the same rat for each data point and a repeated measures one-way ANOVA was used for statistical analysis. (E) Representative electron micrographs of the glomerular capillary wall are shown for (i) diabetes-spiro, and (ii) diabetes-spiro-enzyme samples. Labels indicate endothelial glycocalyx (EnGlx). Bars = 200 nm. TEM quantification at week 8 post-STZ of (F) GEnGlx coverage, and (G) GEnGlx thickness (diabetes-spiro, *n*=4 rats (8 glomeruli); diabetes-spiro-enzyme, *n*=5(14)) confirmed enzyme degradation of GEnGlx. Each triangle, or square on the graph represents a rat. Data are expressed as mean ± SEM. * *P*<0.05; ** *P*<0.01; *** *P*<0.001.

### Increased MMP activity in early DN is ameliorated by MR antagonism

We have previously identified MMP2 and MMP9 as key glycocalyx sheddases.^43,45,46^ Plasma and urine active MMP2 were significantly increased in diabetic rats, with a 1.2-fold increase in plasma (Figure 5A) and a 25-fold increase in urine (Figure 5B). Plasma active MMP9 was not significantly increased (Figure 5C), however urine active MMP9 was significantly increased, with a 14-fold increase compared to controls (Figure 5D). Therapeutic treatment with spironolactone significantly attenuated the diabetes-induced increase in urine MMP2 and MMP9 activities (Figure 5C and 5D), confirming that this MR inhibitor reduces the activity of both MMPs *in vivo*.

**Figure 5.**
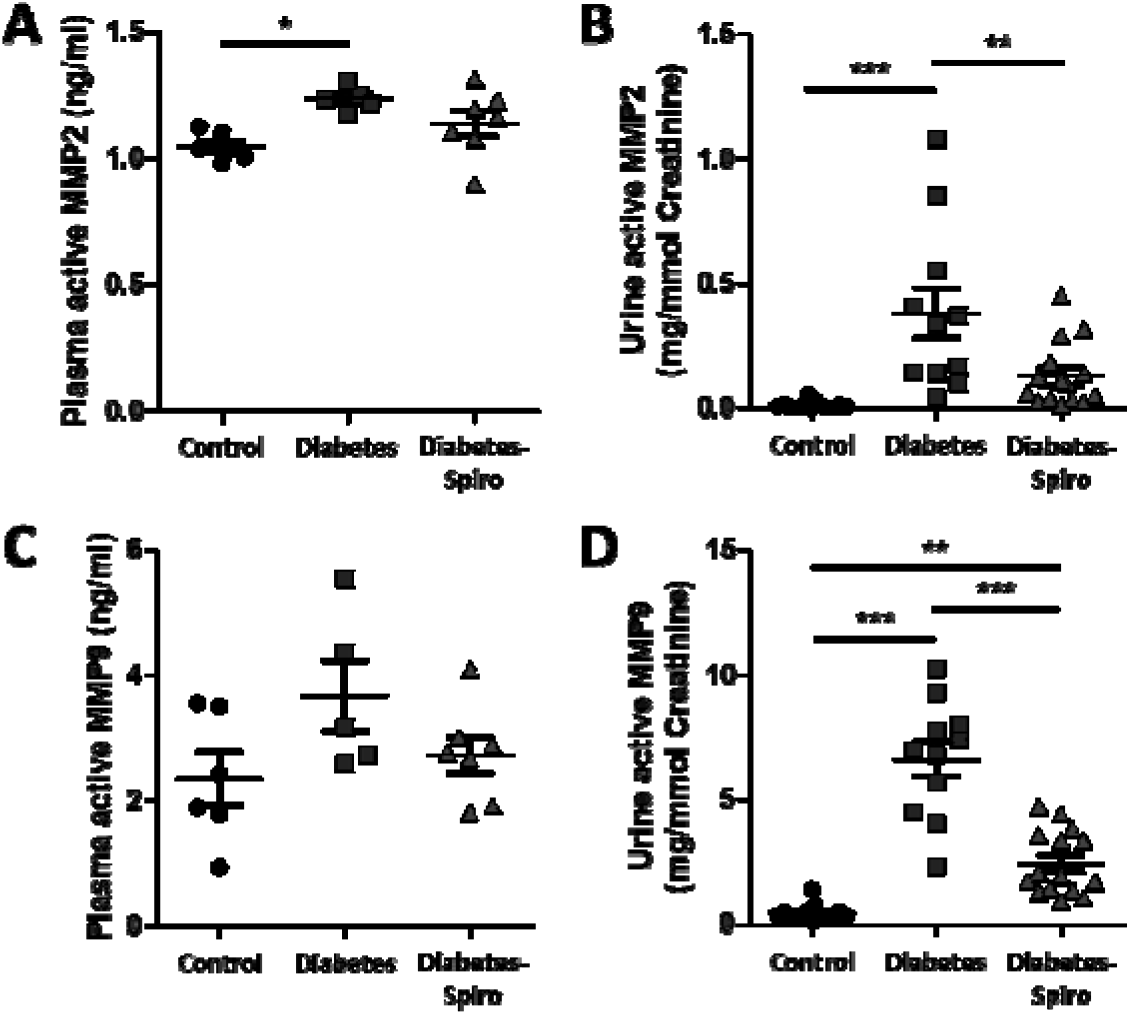
Increased matrix metalloproteinase activity in early diabetic nephropathy is ameliorated by MR antagonism. Matrix metalloproteinase (MMP) activities were measured for the sheddases MMP2 and MMP9. Systemic circulation of (A) plasma active MMP2 and (C) plasma active MMP9 (control, *n*=6; diabetes, *n*=5; diabetes-spironolactone (spiro), *n*=7) were determined. Systemic circulation of (B) urine active MMP2 and (D) urine active MMP9 (control, *n*=13; diabetes, *n*=11; diabetes-spiro, *n*=15) were determined and normalised to urine creatinine at week 8 post-streptozotocin. Each triangle, or square on the graph represents a rat. Data are expressed as mean ± SEM. * *P*<0.05; ** *P*<0.01; *** *P*<0.001.

### The GEnGlx is damaged in human DN

Human renal biopsies were obtained from two different centres (Bristol, UK and Bari, Italy) (Table 1). *Ulex europaeus* agglutinin I (UEA I) lectin has been established as an excellent marker for human endothelial cells^50,51^ and binds specifically to the EnGlx, on the luminal surface of the GEnC (Figure 6A). Representative images show a reduction in UEA I labelling in renal biopsies from DN patients when compared either to TBMN controls or to histologically normal controls. Following extensive validation in our animal model our peak-to-peak measurement technique has allowed us to confirm GEnGlx damage in human diabetes. This may contribute to the disease phenotype seen in human DN. Peak-to-peak measurement of UEA I labelling, with our blinded automated methodology, demonstrated a significant reduction in GEnGlx thickness with DN (Figure 6B). In the Bristol cohort there was a 4.7-fold decrease in GEnGlx thickness in DN patients compared to TBMN controls. This was replicated in the Bari cohort, with a 7.9-fold decrease in GEnGlx thickness in DN patients compared to controls.

**Figure 6.**
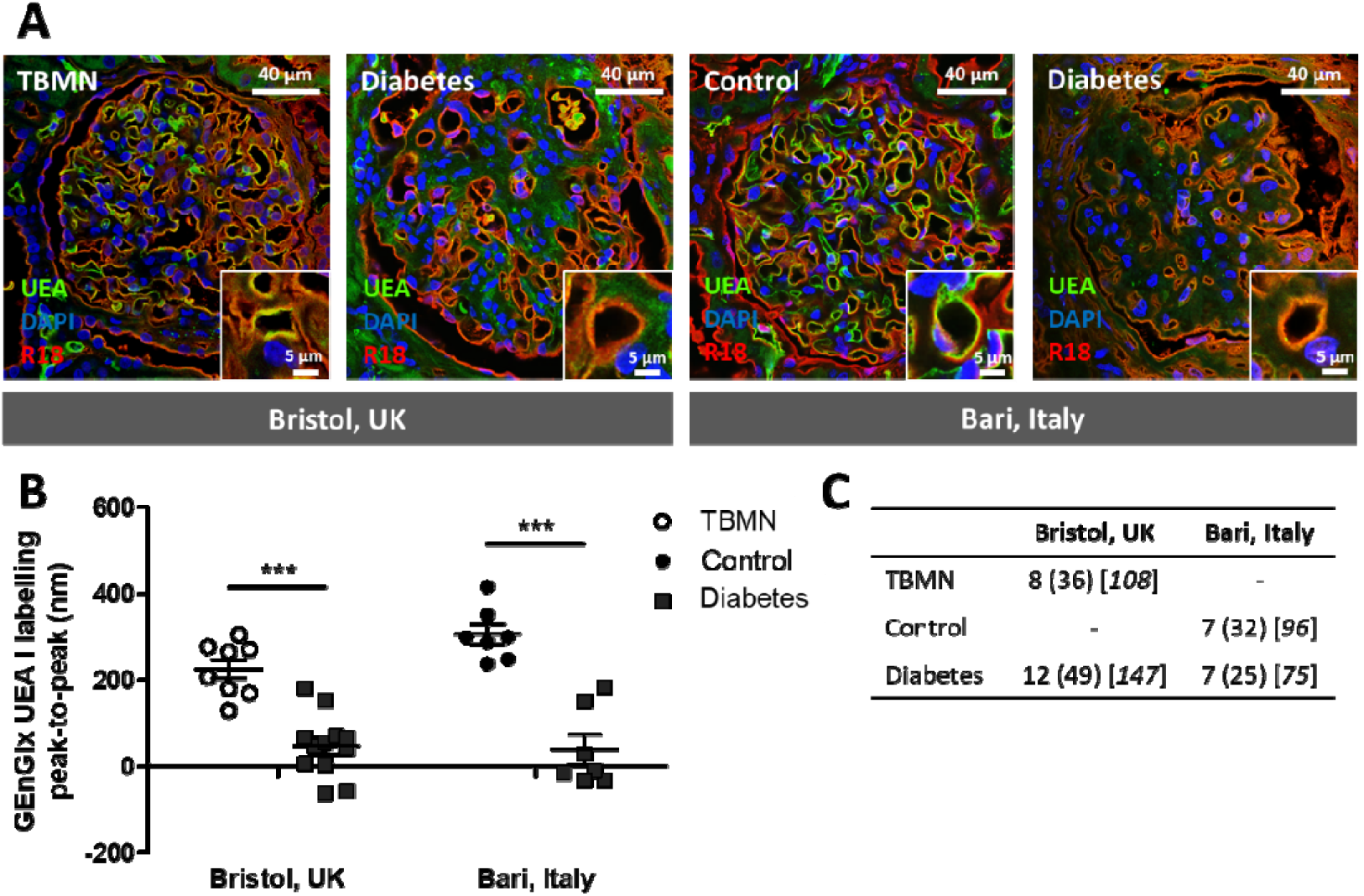
The glomerular endothelial glycocalyx is damaged in human diabetic nephropathy. (A) Representative images show glomerular capillaries labelled red (R18) and the luminal glomerular endothelial glycocalyx (GEnGlx) labelled green with *Ulex europaeus* agglutinin I (UEA-I) in renal biopsies from thin basement membrane nephropathy (TBMN) and diabetic nephropathy (DN) patients from Bristol, UK; and from histologically normal controls and DN patients from Bari, Italy. Bar = 40 _μ_m and 5 _μ_m. (B) Quantification of GEnGlx UEA labelling peak-to-peak (Bristol, UK –TBMN, *n*=8; diabetes, *n*=12; Bari, Italy - control, *n*=7; diabetes, *n*=7) confirms GEnGlx damage may contribute to the disease phenotype seen in human DN. (C) Table indicating the number of samples, glomeruli and capillaries used to analyse each group (*n* = # of samples (# of glomeruli) [*# of capillaries*]. Each dot, or square on the graph represents a patient. Data are expressed as mean ± SEM. *** *P*<0.001.

## Discussion

Using a rat model of diabetes we have demonstrated up-regulation of MMPs, GEnGlx damage and glomerular dysfunction occur early, before other visible markers of DN. MR antagonism in this model preserved the GEnGlx and restored glomerular function. To achieve this, we used a highly sensitive glomerular P*s’*_*alb*_ assay to directly measure glomerular permeability and applied a peak-to-peak measurement technique as an index of glycocalyx thickness. In addition, we have confirmed that GEnGlx damage occurs in human DN and may contribute to the disease phenotype. Together these data suggest that alternative approaches to protect against MR activation-associated GEnGlx damage represent a novel potential therapeutic strategy in patients unable to tolerate direct MR inhibition.

STZ-induced diabetic rats provide a good model of early changes in DN,^26,52,53^ and experiments with pancreatic islet transplantation have shown that albuminuria in this model is due to diabetes rather than STZ toxicity.^54^ As expected, our diabetic rats progressively developed albuminuria. However, urinary albumin excretion is not a sensitive measure of glomerular permeability due to tubular reabsorption of filtered albumin and changes in local haemodynamic factors. Our glomerular P*s’*_*alb*_ assay directly measures glomerular permeability using single capillaries in isolated glomeruli and therefore is not confounded by tubular reuptake of albumin or by haemodynamic factors, including changes in systemic blood pressure.^26^ We observed an increase in glomerular P*s’*_*alb*_ in early DN, and both glomerular permeability and albuminuria were limited by MR antagonism confirming a direct action on the GFB.

The GEnGlx plays an important role in glomerular barrier function, limiting albumin permeability.^18,19,24^ Conventional TEM is often used to directly visualise and measure the EnGlx,^55–57^ however one of the main limitations of TEM is the potential for damage to the EnGlx during tissue preparation.^55,56^ Sample fixation is another factor that can affect quantification of EnGlx, with glycocalyx structure better maintained in perfusion-fixed rather than immersion-fixed tissue.^58^ However, variable tissue perfusion in disease could potentially confound the technique, with a subjective increase in renal perfusion variability noted in diabetic animals. Furthermore, perfusion-fixation techniques are not applicable in humans. Quantification from electron micrographs showed reduced GEnGlx coverage in diabetic rats, restored by inhibiting MR. However, GEnGlx measured by TEM showed only a weak correlation with glomerular permeability. In mice, MOA lectin is known to bind to specific carbohydrate sequences present in the glycocalyx of glomerular endothelium,^43,59,60^ and here we confirmed it bound to the EnGlx on the luminal surface of the R18-labelled GEnC membrane in rats. WGA lectin is known to bind to the EnGlx in rats,^24,49,61^ and with MOA lectin was used to study the GEnGlx. Our peak-to-peak measurement technique has previously been used, *in vivo*^46,49^ and on fixed kidney tissue^43^, as an index of glycocalyx thickness. With peak-to-peak measurement using both MOA and WGA lectin labelling, we have demonstrated that the diabetes-induced reduction in GEnGlx thickness is restored by MR antagonism. In addition, GEnGlx thickness measured by peak-to-peak correlated with glomerular permeability (P*s’*_*alb*_) more strongly than did TEM measurements. This is likely due to perfusion variability and dehydration artifact during tissue processing for TEM. The peak-to-peak measurements using MOA and WGA lectin labelling were also greater than GEnGlx thickness based on TEM. This is likely due to EnGlx collapse during the dehydration procedure that follows fixation in TEM tissue preparation.^55,56,62^ These findings further validate our peak-to-peak measurement technique as a reliable and robust alternative when investigating EnGlx thickness.

The role of the pGlx in diabetes requires further investigation. In our previous work in STZ-induced diabetic mice we found significantly reduced pGlx.^43^ In this study there was a trend toward reduced pGlx but changes were not significant. This may be because the hyperglycaemia-induced morphological changes in the kidneys of STZ-induced diabetic rats are less significant than those observed in STZ-induced diabetic mice.^52^ The hypothesis that endothelial activation can lead to secondary podocyte injury has been supported in some studies.^63–66^ Other glomerular ultrastructural features were not affected. Significantly, STZ-induced diabetic rats did not develop GBM thickening and podocyte effacement by 8 weeks. These data confirm our previous observations that isolated disruption of GEnGlx in early DN is associated with albuminuria and glomerular permeability increases, even in the absence of changes in other glomerular capillary wall components.^26^

The glycocalyx is a complex heterogenous structure. Hyaluronan is a glycocalyx component that has been shown to be shed in diabetes.^67^ In this study we have shown that MR antagonism preserves the GEnGlx, reduces the increased P*s’*_*alb*_ and retards the development of albuminuria in DN. Using enzymatic degradation of the GEnGlx with hyaluronidase in spironolactone-treated diabetic rats we demonstrated that stripping the restored glycocalyx increased albuminuria back to diabetes-induced levels, confirming the importance of GEnGlx preservation in this model of DN. Previously in mice, using a combination of hyaluronidase and chondroitinase we have shown that enzymatic GEnGlx depletion resulted in a significant increase in endothelial permeability.^26,68^ Here, we used hyaluronidase in isolation because it has a short half-life in circulation, 3.2 minutes in rats,^69^ and a molecular weight of 61 kDa thereby limiting its effects to the EnGlx. Consistent with our previous observations,^26,68^ enzyme treatment reduced the GEnGlx, with no effect on pGlx, suggesting enzyme activity was focused within the vessel lumen.

The underlying mechanism of damage in this model appears to be MR-induced MMP induction. We have shown increased plasma and urine MMP2 activity in diabetic rats, and increased MMP9 activity in urine. In humans, plasma MMP2 levels were increased in adults with type 1 and type 2 diabetes.^70–72^ Studies have also found elevated activities of urinary MMP2 and MMP9 in patients with type 1 and type 2 diabetes.^73–75^ Recently in mice, we have demonstrated increased MMP activities induced by excess aldosterone^46^ and by diabetes.^43^ In diabetic mice, we showed both MMP2 and MMP9 activities were enhanced in plasma and urine, with more marked changes noted in urine.^43^ In the current study, changes in plasma MMP9 activity in diabetic rats showed a similar trend to urinary changes, although they did not reach significance. Consistent with our recent findings, the diabetes-induced increase in MMP activity was associated with reduced GEnGlx and an increase in albuminuria. In addition to preserved GEnGlx and normalised glomerular permeability, we have shown that MR antagonism in STZ-induced rats reduced urinary active MMP2 and MMP9, highlighting a mechanism of glycocalyx protection. Similar to urinary changes, there was a trend towards reduced plasma MMP activity with MR antagonism, however changes were not significant. Our findings are consistent with our previously published work that treatment with vascular endothelial growth factor A_165_b, angiopoietin-1 and an MMP2/9 inhibitor reversed damage to the GEnGlx and restored glomerular permeability in early DN.^26,42,43^ In humans, the absence of GBM or podocyte changes in early disease, along with systemic endothelial and glycocalyx dysfunction seen in both type 1^38^ and type 2 diabetes,^39^ strongly implicate GEnGlx damage as a key initiator of albuminuria in DN.^18,19,37^ Using our peak-to-peak technique (UEA I/R18) we have demonstrated that GEnGlx damage occurs in human DN and may contribute to the disease phenotype. This work highlights the importance of work aimed at identifying therapeutic targets capable of limiting EnGlx degradation in disease. Such therapies would protect and restore the glycocalyx and would be of benefit in glomerular diseases, including DN, and also in systemic vascular disease. ^76–78^

In summary, we have shown that MR antagonism prevented damage to the GEnGlx and normalised glomerular albumin permeability in early DN. In addition, critically we have demonstrated that GEnGlx damage may contribute to the disease phenotype in human DN. Alternative approaches to block MR-induced GEnGlx dysfunction warrant further investigation, to reproduce the benefit of MR antagonists in DN without the adverse effects.

## Data Availability

Non sensitive data will be made available to genuine researchers following publication of the original article

## Acknowledgments

The authors would like to thank the Medical Research Council in addition to the Wolfson Foundation for establishing the Wolfson Bioimaging Facility and gratefully acknowledge the Wolfson Bioimaging Facility for their support and assistance in this work. Specifically, Dr Stephen Cross for developing our peak-to-peak automated methodology.

This work was supported by Kidney Research UK grants (RP_031_20180306, S/RP/2015/10, ID_004_20170330 and IN_004_20190305); a Diabetes UK grant (18/0005795); a British Heart Foundation grant (PG/15/81/31740 for K.L.O.); and a Medical Research Council Clinical Research Training Fellowship grant (MR/M018237/1 for M.J.B.).

P.P., L.G. and S.C.S. are members of the Biomarker Enterprise to Attack Diabetic Kidney Disease (BEAt-DKD) consortium and as such this project has received funding from the Innovative Medicines Initiative Joint Undertaking (IMI 2 JU) under grant agreement no. 115974. This Joint Undertaking receives support from the European Union’s Horizon 2020 Research and Innovation Programme and the European Federation of Pharmaceutical Industries and Associations.

## Supplemental Material

This article contains the following supplemental material:

### Supplementary Methods

**Type 1 diabetic model**

**Tissue collection**

**Glomerular albumin permeability (Ps’alb) assay**

**Transmission electron microscopy**

**Lectin staining**

**MMP activity**

### Supplementary References

### Supplementary Figures

**Figure S1**. Streptozotocin (STZ)-induced rats were hyperglycaemic and development of proteinuria in early diabetic nephropathy limited by MR antagonism.

**Figure S2**. Early diabetic nephropathy not significantly associated with other endothelial, podocyte or glomerular parameters.

**Figure S3**. Enzymatic degradation of the endothelial glycocalyx with hyaluronidase is not associated with podocyte glycocalyx damage.

## References

1. UK Renal Registry. UK Renal Registry 22nd Annual Report – data to 31/12/2018, Bristol, UK. Available from http://renal.org/audit-research/annual-report. Published online 2020.

2. NHS Digital. National Diabetes Audit, 2015-16 Report 2a: Complications and Mortality. Available from https://digital.nhs.uk/data-and-information/publications/statistical/national-diabetes-audit. Published online 2017.

3. Strippoli GF, Bonifati C, Craig ME, Navaneethan SD, Craig JC. Angiotensin converting enzyme inhibitors and angiotensin II receptor antagonists for preventing the progression of diabetic kidney disease. Cochrane Database Syst Rev. 2006;2006(4). doi:10.1002/14651858.CD006257

4. Staessen J, Lijnen P, Fagard R, Verschueren LJ, Amery A. Rise in plasma concentration of aldosterone during long-term angiotensin II suppression. J Endocrinol. 1981;91(3):457–465. doi:10.1677/joe.0.0910457

5. Schjoedt KJ. The renin-angiotensin-aldosterone system and its blockade in diabetic nephropathy: main focus on the role of aldosterone. Dan Med Bull. 2011;58(4):B4265.

6. Mavrakanas TA, Gariani K, Martin P-Y. Mineralocorticoid receptor blockade in addition to angiotensin converting enzyme inhibitor or angiotensin II receptor blocker treatment: an emerging paradigm in diabetic nephropathy: a systematic review. Eur J Intern Med. 2014;25(2):173–176. doi:10.1016/j.ejim.2013.11.007

7. Sato A, Hayashi K, Naruse M, Saruta T. Effectiveness of aldosterone blockade in patients with diabetic nephropathy. Hypertens Dallas Tex 1979. 2003;41(1):64–68. doi:10.1161/01.hyp.0000044937.95080.e9

8. Sato A, Hayashi K, Saruta T. Antiproteinuric effects of mineralocorticoid receptor blockade in patients with chronic renal disease. Am J Hypertens. 2005;18(1):44–49. doi:10.1016/j.amjhyper.2004.06.029

9. Sun L-J, Sun Y-N, Shan J-P, Jiang G-R. Effects of mineralocorticoid receptor antagonists on the progression of diabetic nephropathy. J Diabetes Investig. 2017;8(4):609–618. doi:10.1111/jdi.12629

10. Bianchi S, Bigazzi R, Campese VM. Long-term effects of spironolactone on proteinuria and kidney function in patients with chronic kidney disease. Kidney Int. 2006;70(12):2116–2123. doi:10.1038/sj.ki.5001854

11. Navaneethan SD, Nigwekar SU, Sehgal AR, Strippoli GFM. Aldosterone antagonists for preventing the progression of chronic kidney disease: a systematic review and meta-analysis. Clin J Am Soc Nephrol CJASN. 2009;4(3):542–551. doi:10.2215/CJN.04750908

12. Bakris GL, Agarwal R, Anker SD, et al. Effect of Finerenone on Chronic Kidney Disease Outcomes in Type 2 Diabetes. N Engl J Med. 2020;383(23):2219–2229. doi:10.1056/NEJMoa2025845

13. Pitt B, Rossignol P. Mineralocorticoid Receptor Antagonists in Patients With End-Stage Renal Disease on Chronic Hemodialysis. J Am Coll Cardiol. 2014;63(6):537–538. doi:10.1016/j.jacc.2013.09.057

14. Ng KP, Jain P, Gill PS, et al. Results and lessons from the Spironolactone To Prevent Cardiovascular Events in Early Stage Chronic Kidney Disease (STOP-CKD) randomised controlled trial. BMJ Open. 2016;6(2). doi:10.1136/bmjopen-2015-010519

15. Currie G, Taylor AHM, Fujita T, et al. Effect of mineralocorticoid receptor antagonists on proteinuria and progression of chronic kidney disease: a systematic review and meta-analysis. BMC Nephrol. 2016;17(1):127. doi:10.1186/s12882-016-0337-0

16. Satchell SC. The glomerular endothelium emerges as a key player in diabetic nephropathy. Kidney Int. 2012;82(9):949–951. doi:10.1038/ki.2012.258

17. Haraldsson B, Nyström J, Deen WM. Properties of the glomerular barrier and mechanisms of proteinuria. Physiol Rev. 2008;88(2):451–487. doi:10.1152/physrev.00055.2006

18. Salmon AHJ, Satchell SC. Endothelial glycocalyx dysfunction in disease: albuminuria and increased microvascular permeability. J Pathol. 2012;226(4):562–574. doi:10.1002/path.3964

19. Satchell S. The role of the glomerular endothelium in albumin handling. Nat Rev Nephrol. 2013;9(12):717–725. doi:10.1038/nrneph.2013.197

20. Singh A, Fridén V, Dasgupta I, et al. High glucose causes dysfunction of the human glomerular endothelial glycocalyx. Am J Physiol Renal Physiol. 2011;300(1):F40–48. doi:10.1152/ajprenal.00103.2010

21. Singh A, Ramnath RD, Foster RR, et al. Reactive oxygen species modulate the barrier function of the human glomerular endothelial glycocalyx. PloS One. 2013;8(2):e55852. doi:10.1371/journal.pone.0055852

22. Singh A, Satchell SC, Neal CR, McKenzie EA, Tooke JE, Mathieson PW. Glomerular endothelial glycocalyx constitutes a barrier to protein permeability. J Am Soc Nephrol JASN. 2007;18(11):2885–2893. doi:10.1681/ASN.2007010119

23. Jeansson M, Haraldsson B. Morphological and functional evidence for an important role of the endothelial cell glycocalyx in the glomerular barrier. Am J Physiol Renal Physiol. 2006;290(1):F111–116. doi:10.1152/ajprenal.00173.2005

24. Salmon AHJ, Ferguson JK, Burford JL, et al. Loss of the Endothelial Glycocalyx Links Albuminuria and Vascular Dysfunction. J Am Soc Nephrol JASN. 2012;23(8):1339–1350. doi:10.1681/ASN.2012010017

25. Jeansson M, Haraldsson B. Glomerular size and charge selectivity in the mouse after exposure to glucosaminoglycan-degrading enzymes. J Am Soc Nephrol JASN. 2003;14(7):1756–1765. doi:10.1097/01.asn.0000072742.02714.6e

26. Desideri S, Onions KL, Qiu Y, et al. A novel assay provides sensitive measurement of physiologically relevant changes in albumin permeability in isolated human and rodent glomeruli. Kidney Int. 2018;93(5):1086–1097. doi:10.1016/j.kint.2017.12.003

27. Reitsma S, Slaaf DW, Vink H, van Zandvoort Mamj, oude Egbrink Mga. The endothelial glycocalyx: composition, functions, and visualization. Pflugers Arch. 2007;454(3):345–359. doi:10.1007/s00424-007-0212-8

28. Henry CB, Duling BR. Permeation of the luminal capillary glycocalyx is determined by hyaluronan. Am J Physiol. 1999;277(2):H508–514. doi:10.1152/ajpheart.1999.277.2.H508

29. Vink H, Duling BR. Capillary endothelial surface layer selectively reduces plasma solute distribution volume. Am J Physiol Heart Circ Physiol. 2000;278(1):H285–289. doi:10.1152/ajpheart.2000.278.1.H285

30. Davies PF. Flow-mediated endothelial mechanotransduction. Physiol Rev. 1995;75(3):519–560. doi:10.1152/physrev.1995.75.3.519

31. Dewey CF, Bussolari SR, Gimbrone MA, Davies PF. The dynamic response of vascular endothelial cells to fluid shear stress. J Biomech Eng. 1981;103(3):177–185. doi:10.1115/1.3138276

32. Vink H, Duling BR. Identification of distinct luminal domains for macromolecules, erythrocytes, and leukocytes within mammalian capillaries. Circ Res. 1996;79(3):581–589. doi:10.1161/01.res.79.3.581

33. Vink H, Constantinescu AA, Spaan JA. Oxidized lipoproteins degrade the endothelial surface layer!]: implications for platelet-endothelial cell adhesion. Circulation. 2000;101(13):1500–1502. doi:10.1161/01.cir.101.13.1500

34. Yilmaz O, Afsar B, Ortiz A, Kanbay M. The role of endothelial glycocalyx in health and disease. Clin Kidney J. 2019;12(5):611–619. doi:10.1093/ckj/sfz042

35. Weinbaum S, Tarbell JM, Damiano ER. The structure and function of the endothelial glycocalyx layer. Annu Rev Biomed Eng. 2007;9:121–167. doi:10.1146/annurev.bioeng.9.060906.151959

36. Butler MJ, Down CJ, Foster RR, Satchell SC. The Pathological Relevance of Increased Endothelial Glycocalyx Permeability. Am J Pathol. 2020;190(4):742–751. doi:10.1016/j.ajpath.2019.11.015

37. Satchell SC, Tooke JE. What is the mechanism of microalbuminuria in diabetes: a role for the glomerular endothelium? Diabetologia. 2008;51(5):714–725. doi:10.1007/s00125-008-0961-8

38. Nieuwdorp M, Mooij HL, Kroon J, et al. Endothelial glycocalyx damage coincides with microalbuminuria in type 1 diabetes. Diabetes. 2006;55(4):1127–1132. doi:10.2337/diabetes.55.04.06.db05-1619

39. Broekhuizen LN, Lemkes BA, Mooij HL, et al. Effect of sulodexide on endothelial glycocalyx and vascular permeability in patients with type 2 diabetes mellitus. Diabetologia. 2010;53(12):2646–2655. doi:10.1007/s00125-010-1910-x

40. Satoh M, Kobayashi S, Kuwabara A, Tomita N, Sasaki T, Kashihara N. In vivo visualization of glomerular microcirculation and hyperfiltration in streptozotocin-induced diabetic rats. Microcirc N Y N 1994. 2010;17(2):103–112. doi:10.1111/j.1549-8719.2009.00010.x

41. Jeansson M, Granqvist AB, Nyström JS, Haraldsson B. Functional and molecular alterations of the glomerular barrier in long-term diabetes in mice. Diabetologia. 2006;49(9):2200–2209. doi:10.1007/s00125-006-0319-z

42. Oltean S, Qiu Y, Ferguson JK, et al. Vascular Endothelial Growth Factor-A165b Is Protective and Restores Endothelial Glycocalyx in Diabetic Nephropathy. J Am Soc Nephrol JASN. 2015;26(8):1889–1904. doi:10.1681/ASN.2014040350

43. Ramnath RD, Butler MJ, Newman G, et al. Blocking matrix metalloproteinase-mediated syndecan-4 shedding restores the endothelial glycocalyx and glomerular filtration barrier function in early diabetic kidney disease. Kidney Int. 2020;97(5):951–965. doi:10.1016/j.kint.2019.09.035

44. Manon-Jensen T, Itoh Y, Couchman JR. Proteoglycans in health and disease: the multiple roles of syndecan shedding. FEBS J. 2010;277(19):3876–3889. doi:10.1111/j.1742-4658.2010.07798.x

45. Ramnath R, Foster RR, Qiu Y, et al. Matrix metalloproteinase 9-mediated shedding of syndecan 4 in response to tumor necrosis factor α: a contributor to endothelial cell glycocalyx dysfunction. FASEB J Off Publ Fed Am Soc Exp Biol. 2014;28(11):4686–4699. doi:10.1096/fj.14-252221

46. Butler MJ, Ramnath R, Kadoya H, et al. Aldosterone induces albuminuria via matrix metalloproteinase-dependent damage of the endothelial glycocalyx. Kidney Int. 2019;95(1):94–107. doi:10.1016/j.kint.2018.08.024

47. Gregory MC. The clinical features of thin basement membrane nephropathy. Semin Nephrol. 2005;25(3):140–145. doi:10.1016/j.semnephrol.2005.01.004

48. Savige J, Rana K, Tonna S, Buzza M, Dagher H, Wang YY. Thin basement membrane nephropathy. Kidney Int. 2003;64(4):1169–1178. doi:10.1046/j.1523-1755.2003.00234.x

49. Betteridge KB, Arkill KP, Neal CR, et al. Sialic acids regulate microvessel permeability, revealed by novel in vivo studies of endothelial glycocalyx structure and function. J Physiol. 2017;595(15):5015–5035. doi:10.1113/JP274167

50. Holthöfer H, Virtanen I, Kariniemi AL, Hormia M, Linder E, Miettinen A. Ulex europaeus I lectin as a marker for vascular endothelium in human tissues. Lab Investig J Tech Methods Pathol. 1982;47(1):60–66.

51. Miettinen M, Holthofer H, Lehto VP, Miettinen A, Virtanen I. Ulex europaeus I lectin as a marker for tumors derived from endothelial cells. Am J Clin Pathol. 1983;79(1):32–36. doi:10.1093/ajcp/79.1.32

52. Kitada M, Ogura Y, Koya D. Rodent models of diabetic nephropathy: their utility and limitations. Int J Nephrol Renov Dis. 2016;9:279–290. doi:10.2147/IJNRD.S103784

53. Kodera R, Shikata K, Takatsuka T, et al. Dipeptidyl peptidase-4 inhibitor ameliorates early renal injury through its anti-inflammatory action in a rat model of type 1 diabetes. Biochem Biophys Res Commun. 2014;443(3):828–833. doi:10.1016/j.bbrc.2013.12.049

54. Palm F, Ortsäter H, Hansell P, Liss P, Carlsson P-O. Differentiating between effects of streptozotocin per se and subsequent hyperglycemia on renal function and metabolism in the streptozotocin-diabetic rat model. Diabetes Metab Res Rev. 2004;20(6):452–459. doi:10.1002/dmrr.472

55. Kang H, Deng X. The Endothelial Glycocalyx: Visualization and Measurement. J Biomed. 2017;2:120–123. doi:10.7150/jbm.20986

56. Savery MD, Jiang JX, Park PW, Damiano ER. The endothelial glycocalyx in syndecan-1 deficient mice. Microvasc Res. 2013;87:83–91. doi:10.1016/j.mvr.2013.02.001

57. van den Berg BM, Vink H, Spaan JAE. The endothelial glycocalyx protects against myocardial edema. Circ Res. 2003;92(6):592–594. doi:10.1161/01.RES.0000065917.53950.75

58. Chappell D, Jacob M, Paul O, et al. The glycocalyx of the human umbilical vein endothelial cell: an impressive structure ex vivo but not in culture. Circ Res. 2009;104(11):1313–1317. doi:10.1161/CIRCRESAHA.108.187831

59. Haddad G, Zhu LF, Rayner DC, Murray AG. Experimental glomerular endothelial injury in vivo. PloS One. 2013;8(10):e78244. doi:10.1371/journal.pone.0078244

60. Warner RL, Winter HC, Speyer CL, et al. Marasmius oreades lectin induces renal thrombotic microangiopathic lesions. Exp Mol Pathol. 2004;77(2):77–84. doi:10.1016/j.yexmp.2004.04.003

61. Reitsma S, oude Egbrink Mga, Vink H, et al. Endothelial glycocalyx structure in the intact carotid artery: a two-photon laser scanning microscopy study. J Vasc Res. 2011;48(4):297–306. doi:10.1159/000322176

62. Ebong EE, Macaluso FP, Spray DC, Tarbell JM. Imaging the Endothelial Glycocalyx In Vitro by Rapid Freezing/Freeze Substitution Transmission Electron Microscopy. Arterioscler Thromb Vasc Biol. 2011;31(8):1908–1915. doi:10.1161/ATVBAHA.111.225268

63. Daehn I, Casalena G, Zhang T, et al. Endothelial mitochondrial oxidative stress determines podocyte depletion in segmental glomerulosclerosis. J Clin Invest. 2014;124(4):1608–1621. doi:10.1172/JCI71195

64. Sun YBY, Qu X, Zhang X, Caruana G, Bertram JF, Li J. Glomerular endothelial cell injury and damage precedes that of podocytes in adriamycin-induced nephropathy. PloS One. 2013;8(1):e55027. doi:10.1371/journal.pone.0055027

65. Kanetsuna Y, Takahashi K, Nagata M, et al. Deficiency of endothelial nitric-oxide synthase confers susceptibility to diabetic nephropathy in nephropathy-resistant inbred mice. Am J Pathol. 2007;170(5):1473–1484. doi:10.2353/ajpath.2007.060481

66. Rabelink TJ, de Zeeuw D. The glycocalyx--linking albuminuria with renal and cardiovascular disease. Nat Rev Nephrol. 2015;11(11):667–676. doi:10.1038/nrneph.2015.162

67. Dogné S, Flamion B, Caron N. Endothelial Glycocalyx as a Shield Against Diabetic Vascular Complications: Involvement of Hyaluronan and Hyaluronidases. Arterioscler Thromb Vasc Biol. 2018;38(7):1427–1439. doi:10.1161/ATVBAHA.118.310839

68. Onions KL, Gamez M, Buckner NR, et al. VEGFC Reduces Glomerular Albumin Permeability and Protects Against Alterations in VEGF Receptor Expression in Diabetic Nephropathy. Diabetes. 2019;68(1):172–187. doi:10.2337/db18-0045

69. Wolf RA, Chaung LY, O’Hara D, Smith TW, Muller JE. The serum kinetics of bovine testicular hyaluronidase in dogs, rats and humans. J Pharmacol Exp Ther. 1982;222(2):331–337.

70. Derosa G, Avanzini MA, Geroldi D, et al. Matrix metalloproteinase 2 may be a marker of microangiopathy in children and adolescents with type 1 diabetes mellitus. Diabetes Res Clin Pract. 2005;70(2):119–125. doi:10.1016/j.diabres.2005.03.020

71. Lee SW, Song KE, Shin DS, et al. Alterations in peripheral blood levels of TIMP-1, MMP-2, and MMP-9 in patients with type-2 diabetes. Diabetes Res Clin Pract. 2005;69(2):175–179. doi:10.1016/j.diabres.2004.12.010

72. Maxwell PR, Timms PM, Chandran S, Gordon D. Peripheral blood level alterations of TIMP-1, MMP-2 and MMP-9 in patients with type 1 diabetes. Diabet Med J Br Diabet Assoc. 2001;18(10):777–780. doi:10.1046/j.1464-5491.2001.00542.x

73. McKittrick IB, Bogaert Y, Nadeau K, et al. Urinary matrix metalloproteinase activities: biomarkers for plaque angiogenesis and nephropathy in diabetes. Am J Physiol - Ren Physiol. 2011;301(6):F1326–F1333. doi:10.1152/ajprenal.00267.2011

74. Diamant M, Hanemaaijer R, Verheijen JH, Smit JW, Radder JK, Lemkes HH. Elevated matrix metalloproteinase-2 and -9 in urine, but not in serum, are markers of type 1 diabetic nephropathy. Diabet Med J Br Diabet Assoc. 2001;18(5):423–424. doi:10.1046/j.1464-5491.2001.00476-2.x

75. van der Zijl NJ, Hanemaaijer R, Tushuizen ME, et al. Urinary matrix metalloproteinase-8 and -9 activities in type 2 diabetic subjects: A marker of incipient diabetic nephropathy? Clin Biochem. 2010;43(7-8):635–639. doi:10.1016/j.clinbiochem.2010.02.006

76. Drake-Holland AJ, Noble MI. The important new drug target in cardiovascular medicine--the vascular glycocalyx. Cardiovasc Hematol Disord Drug Targets. 2009;9(2):118–123. doi:10.2174/187152909788488708

77. Broekhuizen LN, Mooij HL, Kastelein JJP, Stroes ESG, Vink H, Nieuwdorp M. Endothelial glycocalyx as potential diagnostic and therapeutic target in cardiovascular disease. Curr Opin Lipidol. 2009;20(1):57–62. doi:10.1097/MOL.0b013e328321b587

78. Becker BF, Chappell D, Bruegger D, Annecke T, Jacob M. Therapeutic strategies targeting the endothelial glycocalyx: acute deficits, but great potential. Cardiovasc Res. 2010;87(2):300–310. doi:10.1093/cvr/cvq137

