## Supplementary methods and data for "Early mineralocorticoid receptor antagonism in diabetic nephropathy limits albuminuria by preserving the glomerular endothelial glycocalyx"

Address: Bristol Renal, Dorothy Hodgkin Building, Whitson St, Bristol, BS1 3NY, UK

**Supplementary Methods**

**Type 1 diabetic model**

**Tissue collection**

**Glomerular albumin permeability (Ps’alb) assay**

**Transmission electron microscopy**

**Lectin staining**

**MMP activity**

**Supplementary References**

**Supplementary Figures**

**Figure S1.** Streptozotocin (STZ)-induced rats were hyperglycaemic and development of proteinuria in early diabetic nephropathy limited by MR antagonism.

**Figure S2.** Early diabetic nephropathy not significantly associated with other endothelial, podocyte or glomerular parameters.

**Figure S3.** Enzymatic degradation of the endothelial glycocalyx with hyaluronidase is not associated with podocyte glycocalyx damage.

Supplementary Methods

Type 1 diabetic model

For induction of diabetes, randomised rats were injected i.p. with 50 mg/kg streptozotocin (STZ)(S0130; Sigma-Aldrich, MO, USA) by adding the appropriate volume of the drug at 25 mg/ml in 10 mM sodium citrate, pH 4.5. Comparisons were made with vehicle-treated rats (10 mM sodium citrate, pH 4.5). Glycemia, by tail-tip blood droplet analysis using a glucometer (Accu-Chek Aviva; Roche, Basel, Switzerland), was measured 3 weeks post-STZ and rats with glycemia ≥15 mmol/l were considered diabetic and included in the study.

To confirm whether MR antagonism could preserve the GEnGlx and limit the development of DN, from 4 weeks after STZ administration, randomised diabetic rats were given daily s.c. injections of spironolactone (S3378; Sigma-Aldrich, MO, USA) at 50 mg/kg, made up in corn oil (C8267; Sigma-Aldrich, MO, USA) for 28 days and comparisons were made with vehicle-treated diabetic rats (corn oil). This spironolactone dose has previously been shown to have no effect on blood pressure.^1–4^

To determine the importance of GEnGlx preservation in this model, enzymatic degradation of the GEnGlx with hyaluronidase was used. Briefly, randomised diabetic rats treated with spironolactone were given hyaluronidase (H3506; Sigma-Aldrich, MO, USA), 200 units in 1 ml, via tail vein injection 1 hour before being culled for tissue collection.

Body weight was monitored regularly after STZ and rats were placed on hydrophobic sand to collect urine. Urinary albumin was quantified with a rat albumin ELISA (E111-125, Bethyl Laboratories, TX, USA), and creatinine was measured using an enzymatic spectrophotometric assay (Konelab T-Series 981845; Thermo Fisher Scientific, MA, USA). Urinary albumin:creatinine ratio (uACR) was calculated as previously described.^5^ Rats were culled at 8 weeks post-STZ injection for tissue collection.

Tissue collection

For tissue collection, rats were anaesthetised with isoflurane in 1L/min oxygen. A midline laparotomy was performed, and the abdominal aorta was cannulated with PE-10 tubing (427400; Becton Dickinson, NJ, USA) to flush both kidneys with Ringer solution (NaCl, 132mM; KCl, 4.6mM; MgSO_4_-7H_2_O 1.3mM; CaCl_2_-2H_2_O 2mM; D(+)glucose, 5.5mM; N-2-hydroxyethylpiperazine-N’-2-ethanesulphonic (HEPES) acid, 3.1mM; HEPES sodium salt, 1.9mM, pH 7.4). The left kidney was then removed for lectin staining (1/4, 4% PFA fixed) and glomerular permeability assay (3/4, sieved for glomeruli) as previously.^6^ The right kidney was subsequently perfusion-fixed with a solution containing 2.5% glutaraldehyde, 0.1M cacodylate and 1% Alcian blue for transmission electron microscopy to quantify GEnGlx thickness and coverage.^5,6^

Glomerular albumin permeability (P*s’_alb_*) assay

The glomerular P*s’_alb_* assay was carried out as previously described.^6^ Briefly, Ringer-perfused kidney was sieved in 4% bovine serum albumin (BSA) in Ringer solution. Isolated glomeruli were incubated in 36.5 μg/ml Octadecyl rhodamine B chloride (R18, O246; Thermo Fisher Scientific, MA, USA) for 15 minutes, then washed in 4% Ringer BSA to remove unbound R18 followed by 15 minutes’ incubation in 30 μg/ml Alexa Fluor 488-BSA (A13100; Thermo Fisher Scientific, MA, USA). An individual glomerulus was trapped on a custom-made petri dish and the perfusate was switched from 30 μg/ml labelled 488-BSA to 30 μg/ml unlabelled BSA. A Nikon Ti-E inverted confocal microscope (Nikon Instruments Inc., NY, USA) was used to capture the fluorescence intensity. The rate of decline in fluorescence intensity within the loop of the capillaries for the first minute was used to calculate P*s’_alb_* as previously described.^6^ Observers were blinded to sample identity.

Transmission electron microscopy

Perfusion-fixed right kidneys were used for transmission electron microscopy (TEM), as previously, to measure GEnC, GBM and podocyte parameters to identify effects on GFB ultrastructure that could explain changes in glomerular permeability.^5,6^ Electron micrographs were taken using a Technai 12 electron microscope (FEI, OR, USA) and image analysis was carried out using established protocols in 3-4 capillary loops per glomerulus and 2–3 glomeruli per animal.^5,6^ Briefly, ImageJ software (SciJava software ecosystem, MD, USA) was used to overlay a grid onto the electron micrograph. The anatomical distance from the luminal phospholipid at sequential grid intersections to the furthest point of glycocalyx was measured as glycocalyx thickness. A glycocalyx thickness ≤ 10nm was considered uncovered and this was expressed as a percentage of total measurements taken on a grid section. GBM width, podocyte foot process width and slit diaphragm width were also measured. Fenestration density and podocyte foot process density were measured by counting the number of each and dividing by the length of GBM used for analysis. Observers were blinded to sample identity.

Lectin staining

*Marasmium oreades* agglutinin (MOA), wheat germ agglutinin (WGA) and *Ulex europaeus* agglutinin I (UEA I) lectins have binding specificities for carbohydrate sequences present in the glycocalyx. MOA lectin binds to non-reducing terminal Galα(1,3)Gal-carbohydrates, WGA lectin binds to the sialyloligosaccharides, N-Acetylglucosamine and N-Acetylneuraminic acid, and UEA I binds to α-L-fucose. Lectins were labelled with biotin or Fluorescein isothiocyanate (FlTC): biotinylated MOA (2 mg/ml; 1:100); FITC-WGA (GTX01502; GeneTex, CA,USA; 5 mg/ml; 1:500); and biotinylated UEA I (GTX01511; GeneTex, CA,USA; 2 mg/ml; 1:200):

Paraffin-embedded kidney sections (5 μm) were dewaxed in Histo-Clear II (National Diagnostics, NC, USA) followed by rehydration in graded ethanol and a wash in PBS. All sections were incubated in blocking buffer (1% BSA in PBS containing 0.1% Tween) for 30 minutes. For biotinylated lectins, this was followed by endogenous biotin blocking using a streptavidin/biotin blocking kit (SP-2002; Vector Laboratories, CA, USA). After 2 washes, the sections were incubated with the biotinylated lectin, pH 6.8, overnight at 4 °C. Buffer only was used as a negative control. After 4 washes, the sections were incubated with streptavidin-Alexa Fluor 488 (1:500, S32354; Thermo Fisher Scientific, MA, USA), pH 6.8, for 1 hour at room temperature. For FITC-labelled lectins, the sections were incubated overnight at 4 °C, pH 6.8. Then for all sections the nuclei were counterstained with 4′,6-diamidino-2-phenylindole (D1306; Thermo Fisher Scientific, MA, USA) and the cell membrane labelled with R18 (1:1000, O246; Thermo Fisher Scientific, MA, USA) were incubated for 10 minutes. After a 2-minute wash in PBS, the coverslips were mounted in Vectashield mounting medium (H-1000; Vector Laboratories, CA, USA) and examined using either an AF600 LX wide-field fluorescence microscope (Leica Microsystems, Milton Keynes, UK) or a Leica SP5-II confocal laser scanning microscope attached to a Leica DMI 6000 inverted epifluorescence microscope.

MMP activity

MMP2 activity was studied using the MMP2 Biotrack activity assay (RPN2631; GE Healthcare, Buckinghamshire, UK), and MMP9 activity was studied using the SensoLyte Plus 520 MMP9 assay (AS-72017; AnaSpec, CA, USA). The manufacturer’s instructions were followed in full. Urinary active MMP2 and -9 were normalised to creatinine.

Supplementary References

1.	Mayyas F, Alzoubi KH, Bonyan R. The role of spironolactone on myocardial oxidative stress in rat model of streptozotocin-induced diabetes. *Cardiovascular Therapeutics*. 2017;35(2):e12242. doi:https://doi.org/10.1111/1755-5922.12242

2.	Banki NF, Ver A, Wagner LJ, et al. Aldosterone Antagonists in Monotherapy Are Protective against Streptozotocin-Induced Diabetic Nephropathy in Rats. *PLOS ONE*. 2012;7(6):e39938. doi:10.1371/journal.pone.0039938

3.	Fujisawa G, Okada K, Muto S, et al. Spironolactone prevents early renal injury in streptozotocin-induced diabetic rats. *Kidney International*. 2004;66(4):1493-1502. doi:10.1111/j.1523-1755.2004.00913.x

4.	Toyonaga J, Tsuruya K, Ikeda H, et al. Spironolactone inhibits hyperglycemia-induced podocyte injury by attenuating ROS production. *Nephrology Dialysis Transplantation*. 2011;26(8):2475-2484. doi:10.1093/ndt/gfq750

5.	Oltean S, Qiu Y, Ferguson JK, et al. Vascular Endothelial Growth Factor-A165b Is Protective and Restores Endothelial Glycocalyx in Diabetic Nephropathy. *J Am Soc Nephrol*. 2015;26(8):1889-1904. doi:10.1681/ASN.2014040350

6.	Desideri S, Onions KL, Qiu Y, et al. A novel assay provides sensitive measurement of physiologically relevant changes in albumin permeability in isolated human and rodent glomeruli. *Kidney Int*. 2018;93(5):1086-1097. doi:10.1016/j.kint.2017.12.003

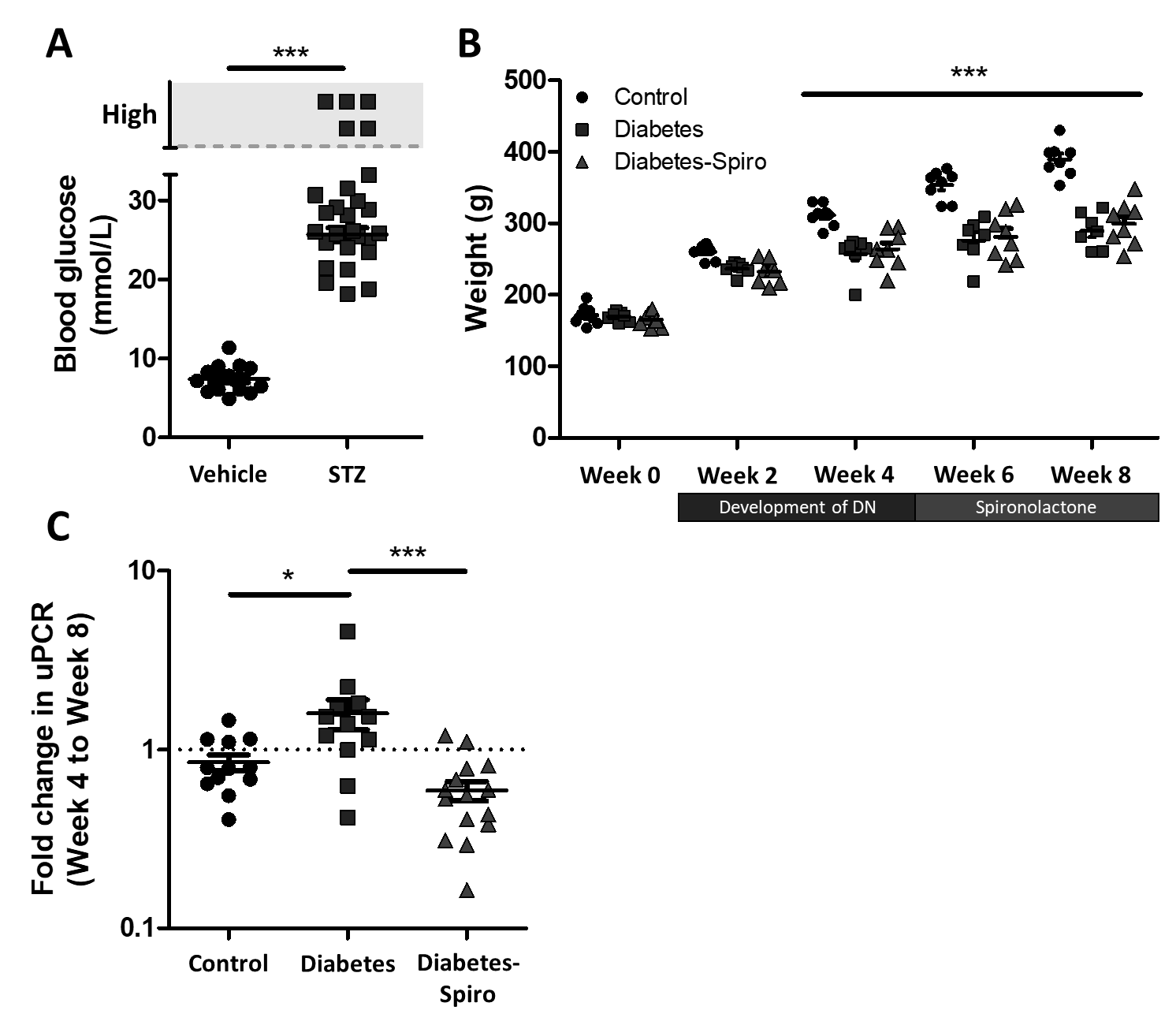

**Figure S1. Streptozotocin (STZ)-induced rats were hyperglycaemic and development of proteinuria in early diabetic nephropathy limited by MR antagonism.** (A) Rats were hyperglycaemic at week 3 post-STZ (vehicle, *n*=14; STZ, *n*=27). (B) Body weight was not significant between groups when compared at week 0 but from week 4 post-STZ both vehicle and spironolactone (spiro) treated diabetic rats were significantly lower than control rats (control, *n*=8; diabetes, *n*=7; diabetes-spiro, *n*=8). (C) Urinary protein:creatinine ratio (uPCR) was determined at week 4 and week 8 showing treatment with spiro reduced the fold change in uPCR from initiation of treatment (control, *n*=12; diabetes, *n*=12; diabetes-spiro, *n*=15). A base-10 log scale is used for the Y axis. Each dot, triangle, or square on the graph represents a rat. Data are expressed as mean ± SEM. * *P*<0.05; *** *P*<0.001.

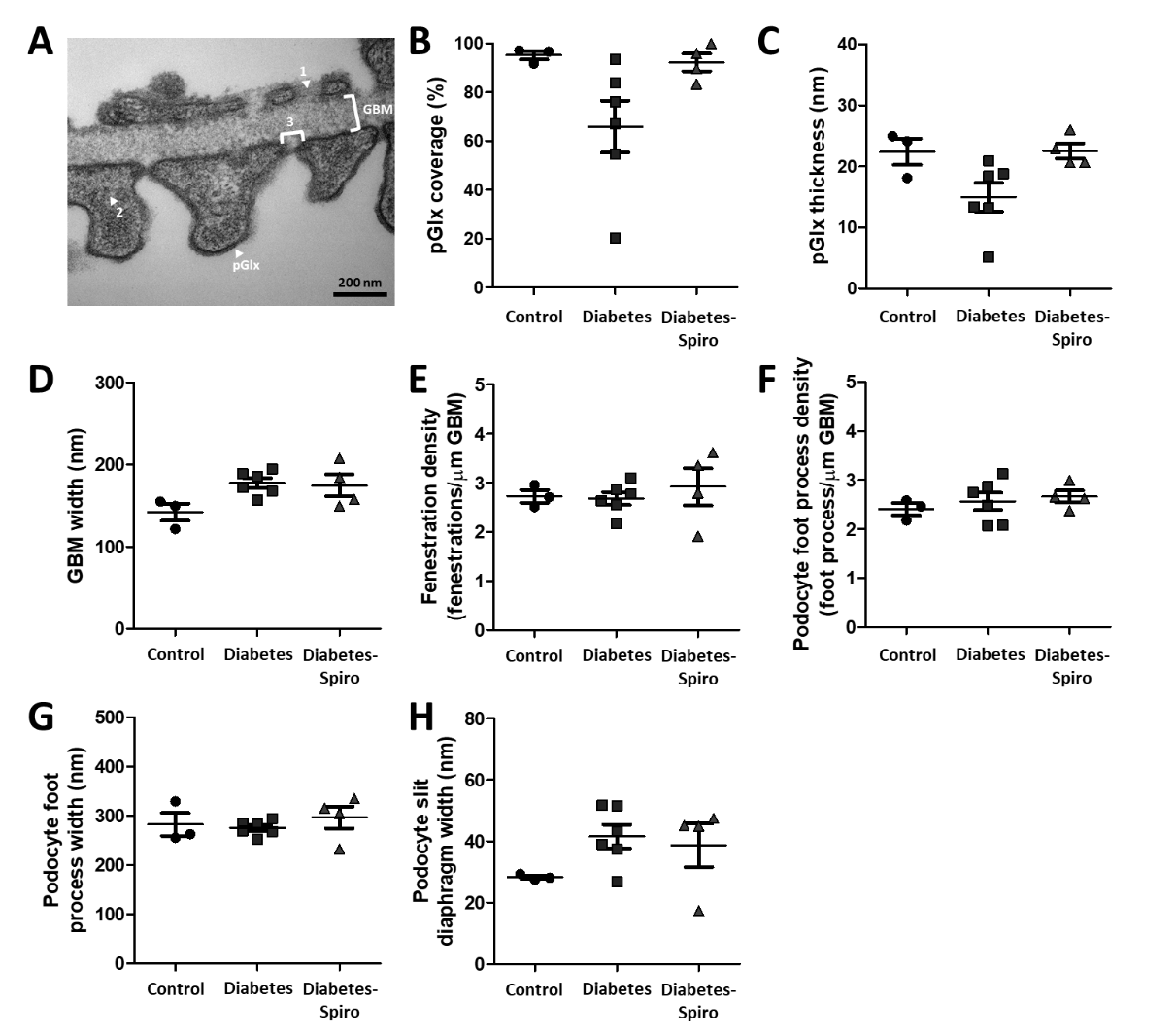

**Figure S2. Early diabetic nephropathy not significantly associated with other endothelial, podocyte or glomerular parameters.** Rats were perfusion-fixed for transmission electron microscopy with cacodylate buffer containing glutaraldehyde and Alcian blue. (A) Representative electron micrographs of the glomerular capillary wall with labels indicating podocyte glycocalyx (pGlx), glomerular basement membrane (GBM), (1) fenestration, (2) podocyte foot process, and (3) slit diaphragm. Bar = 200 nm. Quantification at week 8 post-STZ of (B) pGlx coverage, (C) pGlx thickness, (D) GBM width, (E) fenestration density, (F) podocyte foot process density, (G) podocyte foot process width, and (H) slit diaphragm width (control, *n*=3 rats (9 glomeruli); diabetes, *n*=6(13); diabetes-spiro, *n*=4(8)). Each dot, triangle, or square on the graph represents a rat. Data are expressed as mean ± SEM.

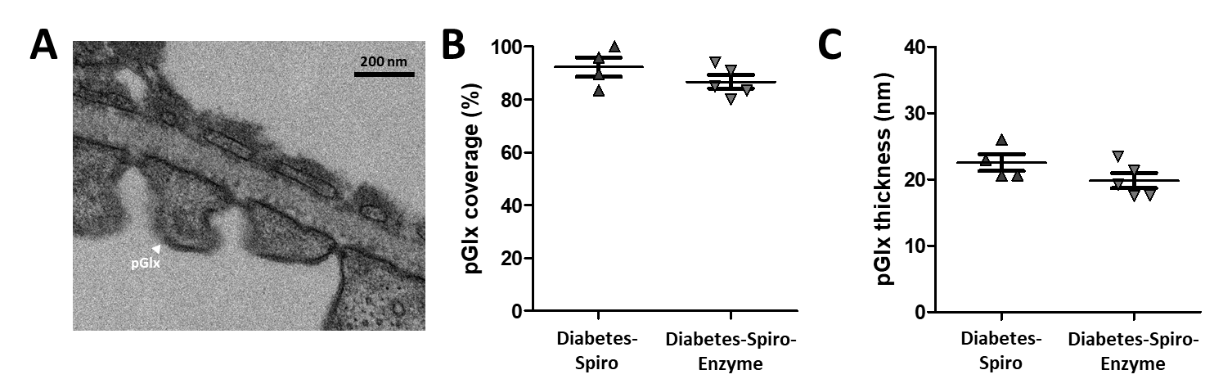

**Figure S3.** **Enzymatic degradation of the endothelial glycocalyx with hyaluronidase is not associated with podocyte glycocalyx damage.** Rats were perfusion-fixed for transmission electron microscopy with cacodylate buffer containing glutaraldehyde and Alcian blue. (A) Representative electron micrograph of the glomerular capillary wall with a label indicating podocyte glycocalyx (pGlx). Bar = 200 nm. Quantification at week 8 post-STZ of (B) pGlx coverage, and (C) pGlx thickness confirmed enzyme degradation had no effect on the pGlx suggesting hyaluronidase activity was focused within the circulation (diabetes-spironolactone (spiro), *n*=4 rats (8 glomeruli); diabetes-spiro-enzyme, *n*=5(14)). Each dot, triangle, or square on the graph represents a rat. Data are expressed as mean ± SEM.
